# Linezolid population pharmacokinetic model in plasma and cerebrospinal fluid among patients with tuberculosis meningitis

**DOI:** 10.1101/2023.04.06.23288230

**Authors:** Noha Abdelgawad, Sean Wasserman, Mahmoud Tareq Abdelwahab, Angharad Davis, Cari Stek, Lubbe Wiesner, John Black, Graeme Meintjes, Robert J. Wilkinson, Paolo Denti

## Abstract

**Background:** Linezolid is being evaluated in novel treatment regimens for tuberculous meningitis (TBM). The pharmacokinetics of linezolid have not been characterized in this population, particularly in cerebrospinal fluid (CSF) where exposures may be affected by changes in protein concentration and rifampicin co-administration.

**Methods:** This was a sub-study of a phase 2 clinical trial of intensified antibiotic therapy for adults with HIV-associated TBM. Participants in the intervention groups received high-dose rifampicin (35 mg/kg) plus linezolid 1200 mg daily for 28 days followed by 600 mg daily until day 56. Plasma was intensively sampled, and lumbar CSF was collected at a single timepoint in a randomly allocated sampling window, within 3 days after enrolment. Sparse plasma and CSF samples were also obtained on day 28. Linezolid concentrations were analyzed using non-linear mixed effects modelling.

**Results:** 30 participants contributed 247 plasma and 28 CSF linezolid observations. Plasma PK was best described by a one-compartment model with first-order absorption and saturable elimination. The typical value of maximal clearance was 7.25 L/h. Duration of rifampicin co-treatment (compared on day 3 versus day 28) did not affect linezolid pharmacokinetics. Partitioning between plasma and CSF correlated with CSF total protein concentration up to 1.2 g/L where the partition coefficient reached a maximal value of 37%. The equilibration half-life between plasma and CSF was estimated at ∼3.5 hours.

**Conclusion:** Linezolid was readily detected in CSF despite co-administration of the potent inducer rifampicin at high doses. These findings support continued clinical evaluation of linezolid plus high-dose rifampicin for the treatment of TBM in adults.

## INTRODUCTION

Tuberculous meningitis (TBM) is the most fatal and debilitating form of tuberculosis with a particularly high burden among people living with HIV [1]. One reason for severe outcomes is that the current treatment regimen for TBM is based on treatment for pulmonary TB, and may result in suboptimal central nervous system (CNS) concentrations [2]. Drugs targeted at TBM should cross several barriers to reach the site of disease, including the blood-brain barrier (BBB) and the blood-cerebrospinal fluid barrier (BCSFB) that separate the systemic circulation from their site of action in the CNS. These barriers pose a therapeutic challenge by limiting entry of drugs into the CNS. Moreover, disease-related changes in BBB permeability and dynamic changes in protein concentrations may have important implications for drug penetration into the brain [3].

Linezolid, an oxazolidinone antibiotic, is highly effective for the treatment of drug-resistant pulmonary TB. Linezolid is also used to treat Gram-positive bacterial infections in the CNS [4–6], where good drug penetration has been documented, making it an attractive candidate for TBM treatment [7–9]. Small observational studies have shown improved clinical parameters with linezolid use in children and adults with TBM [10,11]. Based on these encouraging observations, linezolid is being investigated as part of intensified antibiotic therapy in several clinical trials for TBM [12].

Specific features of TBM may influence the pharmacokinetics (PK) of linezolid, with potential implications for safety and efficacy, given its narrow therapeutic window. These include host factors (such as body size) and disease factors, including CSF protein concentrations and BBB permeability. Also, clinical trials provide linezolid along with high-dose rifampicin in TBM treatment regimens. As a potent inducer of the cytochrome P450 system and upregulator of drug transporters [13], rifampicin could potentially affect the PK of linezolid. Studies in healthy volunteers and pulmonary TB have shown a moderate reduction in linezolid exposure when co-administered with standard dose rifampicin [14,15]. The impact on site of disease (CSF) concentrations and clinical implications of this pharmacokinetic interaction is unknown but could theoretically lead to suboptimal treatment or the development of antimicrobial resistance.

The objectives of this analysis were to describe the PK of linezolid in plasma and CSF of adults with TBM, to explore the effect of high-dose rifampicin on linezolid PK, evaluate covariate effects on plasma and CSF drug levels, and simulate exposures for optimized dosing strategies.

## METHODOLOGY

### Study data

This was a sub-study of LASER-TBM [16], a phase IIb, open-label trial that evaluated safety and PK of intensified antibiotic therapy in adults with HIV and TBM [12]. Participants were enrolled from 4 public hospitals in Cape Town and Gqeberha, South Africa, and randomized to study interventions within 5 days of starting antituberculosis treatment. The standard of care (control) group received fixed-dose combination oral tablets (rifampicin 10 mg/kg, isoniazid 5 mg/kg, pyrazinamide 25 mg/kg, and ethambutol 15 mg/kg) according to World Health Organization (WHO) weight bands. Participants allocated to experimental groups were administered the standard regimen with a higher dose of rifampicin (35 mg/kg in total, using bespoke weight bands [17]) and linezolid for 56 days (1200 mg once daily for the first 28 days, then reduced to 600 mg once daily) with or without aspirin. All participants received adjunctive dexamethasone.

Pharmacokinetic sampling visits were scheduled on Day 3 (±2 days) and Day 28 (±2 days) after study entry. At the Day 3 visit, plasma was collected at pre-dose, 0.5, 1, 2, 3, 6, 8-10, and 24 hours post-dose (intensive) and on Day 28 at pre-dose, 2-, and 4-hours post-dose (sparse). Sparse sampling was performed on Day 3 for participants who declined intensive sampling or for whom intensive sampling could not be done. One lumbar CSF sample was collected at each pharmacokinetic sampling visit, with sample timing randomized to intervals of 1-3, 3-6, 6-10, and 24 hours after dosing. Immediately following collection, samples were processed directly and then stored at -80°C. Clinical information was collected, and full blood count and serum chemistry were obtained at each visit. Total protein, albumin, and glucose were measured in CSF samples.

Linezolid plasma and CSF concentrations were measured in the Division of Clinical Pharmacology at the University of Cape Town. The plasma assay summary has been described previously [18]. CSF samples were processed with a protein precipitation extraction method using linezolid-d3 as the internal standard, followed by high-performance liquid chromatography with tandem mass spectrometry detection (LC-MS/MS). Cholesterol and 4-beta hydroxy cholesterol (4β-OHC) were also measured in pre-dose plasma samples collected on both PK visits. 4β-OHC was also measured with an LC-MS/MS assay in the Division of Clinical Pharmacology at the University of Cape Town. Cholesterol plasma concentrations were measured at the South African National Health Laboratory using standard methodology. 4β-OHC is a metabolite of cholesterol formed by CYP3A4 and the ratio between its concentration and that of cholesterol is used a marker of CYP3A4/5 endogenous activity [19].

Informed consent was obtained from all participants or their proxies. The study was approved by the University of Cape Town Human Research Ethics Committee (UCT HREC reference: 293/2018), Walter Sisulu University (HREC reference: 012/2019), and the South African Health Products Regulatory Authority (reference number 20180622). The trial is registered on clinicaltrials.gov (NCT03927313).

### Pharmacokinetic modelling

Nonlinear mixed-effects modelling was used to create a population PK model describing linezolid PK in both plasma and lumbar CSF. The model was developed sequentially; first describing plasma linezolid and then including CSF concentrations.

For the plasma PK, we tested one- and two-compartment disposition models with linear or saturable elimination and first-pass effect. Lag time and transit compartments were tested to capture the delay in the absorption process. Allometric scaling of clearance and volume parameters was tested as per Anderson and Holford [20] using the fixed power exponents of 0.75 for clearance and 1 for volume and either total body weight or fat-free mass (FFM) (calculated based on the formula in Janmahasatian *et al*. [21]) as body size descriptors. The CSF concentrations were described using a hypothetical effect compartment linked to the central (plasma) compartment, which estimates the first-order equilibration rate constant of linezolid between the central and the effect compartments (*k*_*plasma*−*CSF*_) and the pseudo-partition coefficient (*PPC*). Further details on modelling approach are available in the supplementary file. Between-subject, between-visit and between-occasion variabilities were considered on various PK parameters. Each PK sampling day (day 3 and day 28) was considered as a separate visit. Each dose and its following samples were considered a separate occasion, therefore, the dose before the sampling visit along with the predose concentration were treated as a separate occasion from the dose administered during the PK visit and the following concentrations. Residual unexplained variability was best described by a combined additive and proportional error model. Censored plasma values that were below the limit of quantification (BLQ) were imputed to half the lower limit of quantification (LLOQ) and their additive error component inflated by LLOQ/2 [22].

Following the development of the structural model, we tested the effect of potential covariates including creatinine clearance (calculated with the Cockcroft-Gault equation [23]), age, study visit, duration of concomitant rifampicin treatment, study site, and treatment arm. For the CSF PK parameters *PPC* and *k*_*plasma*−*CSF*_, we also tested the effect of CSF total protein, albumin, and glucose concentrations. The precision of the parameter estimates of the final model, expressed as 95% confidence intervals, was assessed using sampling importance resampling (SIR) [24].

### Simulations

The model-derived area under the concentration-time curve from time 0 to 24 hours post-dose (AUC_0-24h_) and the concentration at 24 hours post-dose (C_24h_) were calculated for the available profiles. Monte Carlo simulations (n=10,000) were performed using final model parameters to simulate concentration-time profiles in plasma and CSF following daily linezolid doses of 600 mg or 1200 mg at steady-state for a typical participant with median FFM of 45 kg and CSF protein of 0.995 mg/mL.

## RESULTS

### Study data

Thirty participants underwent PK sampling on the first PK visit on day 3 of the study and 18 participants had PK sampling at the second PK visit on day 28, one of whom was excluded from this analysis because all 3 samples were BLQ (later confirmed to have missed dosing). Reasons for missing the second PK visit included death, interrupting linezolid dose due to adverse events, or withdrawing consent. There were 247 plasma concentrations (6 were BLQ) and 28 CSF concentrations (7 were BLQ) available for PK modeling. All participants were receiving linezolid 1200 mg daily at the first PK visit; on day 28, 13 received 1200 mg and 4 received 600 mg. Baseline clinical characteristics are summarized in **Table 1**. Median duration on rifampicin therapy was 5 days (range 0 – 7) at the Day 3 PK visit and 30 days (range 27 – 38) at the Day 28 visit. Median CSF total protein concentrations decreased from 1.46 g/L (range 0.31 – 54.7) at Day 3 to 0.75 g/L (range 0.22 – 2.19) at Day 28.

**Table 1.**
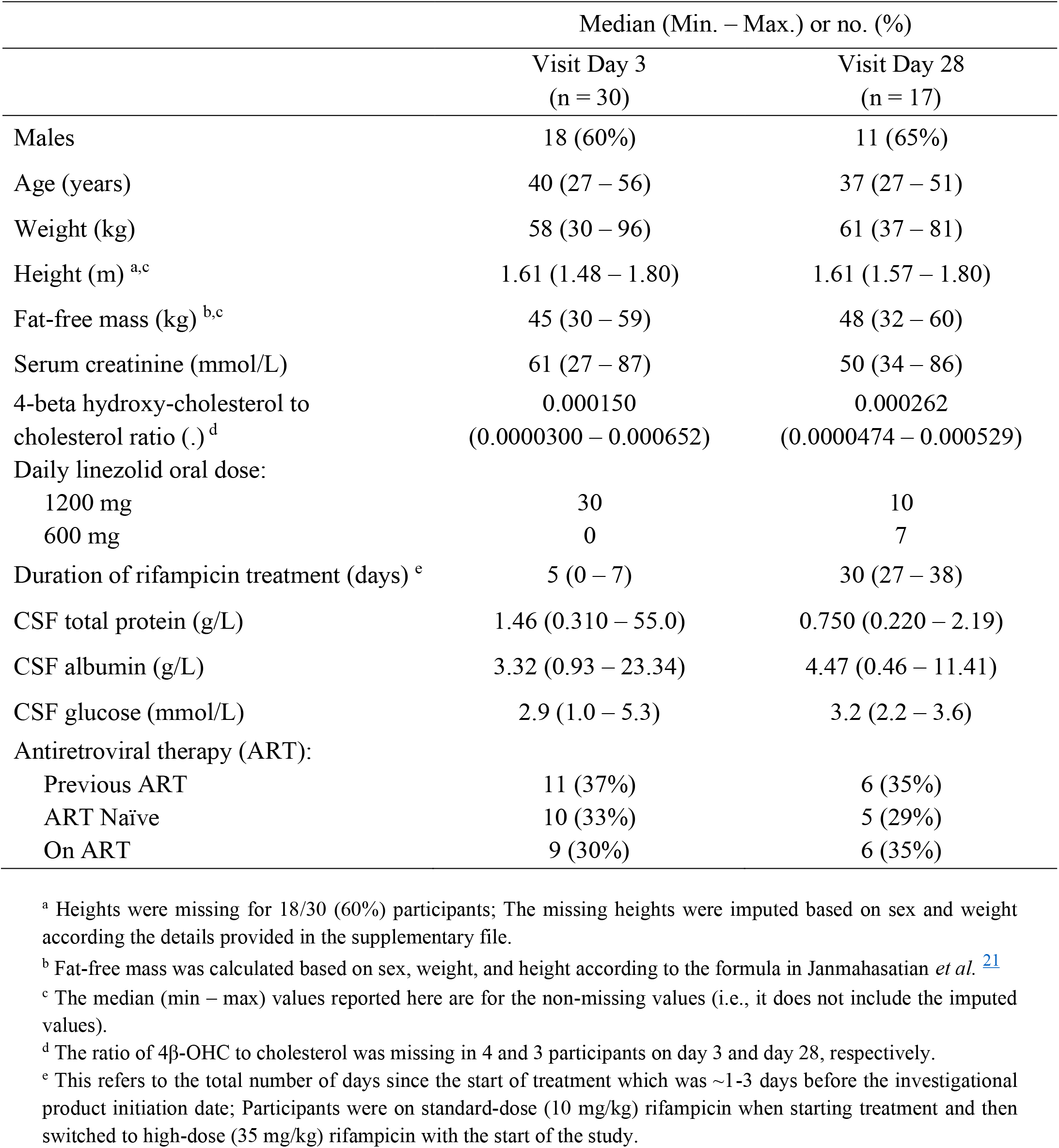
Clinical characteristics

### Pharmacokinetic modelling

The plasma PK of linezolid was best characterized by a one-compartment disposition model, saturable elimination with Michaelis–Menten, and first-order absorption preceded by a chain of transit compartments. A schematic diagram of the model in shown in **Figure 1**. Two-compartment disposition was tested but did not result in a significant improvement of fit. Maximal clearance (*CLmax*) and volume of distribution (*V*) were allometrically scaled using FFM (drop in objective function value (dOFV) = -30, compared to dOFV = -7.7 when using total body weight). In a typical participant (median FFM 45 kg) the value of maximal clearance (*CLmax*) was 6.78 L/h, the Michaelis-Menten constant (*km*), which is a parameter that governs saturable hepatic elimination and represents the linezolid concentration at which half the *CLmax* is reached, was 33.0 mg/L, and the volume of distribution (*V*) was 40.7 L. The inclusion of between-visit variability in *CLmax* improved the model fit, but no systematic increase or decrease with time on treatment was observed. Longitudinal changes in clearance were explored by testing auto-inhibition and duration of rifampicin co-treatment, but no significant effect was found for either. We also could not find any effect when testing the ratio of 4β-OHC to cholesterol, creatinine clearance, or age on *CLmax* and bioavailability (*F*). The final parameter estimates are presented in **Table 2**. A visual predictive check showing adequate model fit is depicted in **Figure S1** in the supplementary file.

**Table 2.** Final population pharmacokinetic parameter estimates for linezolid in plasma and lumbar cerebrospinal fluid

| Parameter | Estimate<br>(95% confidence interval) <sup>a</sup> |
| --- | --- |
| Maximal clearance, $CL_{max}$ (L/h) <sup>b</sup> | 7.25 (6.09 – 8.86) |
| Michaelis-Menten constant, $km$ (mg/L) | 27.2 (16.0 – 46.4) |
| Volume of distribution, $V$ (L) <sup>b</sup> | 40.8 (37.9 – 43.6) |
| Bioavailability, $F$ (.) | 1 Fixed |
| Mean transit time, $MTT$ (h) | 0.211 (0.112 – 0.342) |
| No. of absorption transit compartments, $NN$ (.) | 5.68 (2.36 – 11.8) |
| Absorption rate constant, $ka$ (h<sup>-1</sup>) | 1.21 (0.831 – 1.76) |
| Proportional error plasma (%) | 21.5 (18.8 – 24.7) |
| Additive error plasma (mg/L) <sup>d</sup> | 0.173 (0.0379 – 0.355) |
| Between-subject variability (BSV) in $CL_{max}$ (%) | 9.60 (3.44 – 13.9) |
| Between-visit variability (BVV) in $CL_{max}$ (%) | 20.3 (15.3 – 26.9) |
| Between-occasion variability (BOV) in $ka$ (%) | 87.9 (66.4 – 110) |
| BOV in $MTT$ (%) | 110 (75.8 - 144) |
| Equilibration rate constant to CSF, $k_{plasma-CSF}$ (h<sup>-1</sup>) <sup>e</sup> | 0.198 (0.0849 – 0.340) |
| Maximal pseudo-partition coefficient to CSF, $PPC_{max}$ (.) | 0.365 (0.238 – 0.566) |
| CSF protein at which $PPC_{max}$ is reached, $CSF\ protein_{max}$ (g/L) <sup>f</sup> | 1.18 (0.730 – 1.90) |
| Proportional error CSF (%) | 91.5 (63.3 – 151) |
| Additive error CSF (mg/L) <sup>d</sup> | 0.02 Fixed |

<sup>a</sup> Values in parentheses are the 95% confidence interval, computed with sampling importance resampling (SIR) on the final model.
<sup>b</sup> The values of $CL_{max}$ and $V$ were allometrically scaled, so the typical values reported here refer to the typical participant, i.e. a median FFM of 45 kg.
<sup>d</sup> The estimate of the additive component of the error was not significantly different from its lower boundary of 20% of LLOQ, so it was fixed to this value.
<sup>e</sup> Corresponds to an equilibration half-life of 3.45 (2.13 - 7.33) h
<sup>f</sup> For $CSF\ protein < CSF\ protein_{max}$ (i.e., the breakpoint): $PPC_i = PPC_{max} \cdot (slope \cdot (CSF\ protein - breakpoint))$ , where the *breakpoint* was estimated to be 1.18 mg/mL, and the *slope* was calculated to be 0.847 from the following equation: $slope = (amplitude - intercept) / (breakpoint - 0)$ , where the *intercept* and *amplitude* were fixed to 0 and 1, respectively. For $CSF\ protein \geq CSF\ protein_{max}$ : $PPC_i = PPC_{max}$ . The *PPC-CSF* protein relationship is depicted in **Figure 2**, and more details are provided in the supplementary file.

**Figure 1:**
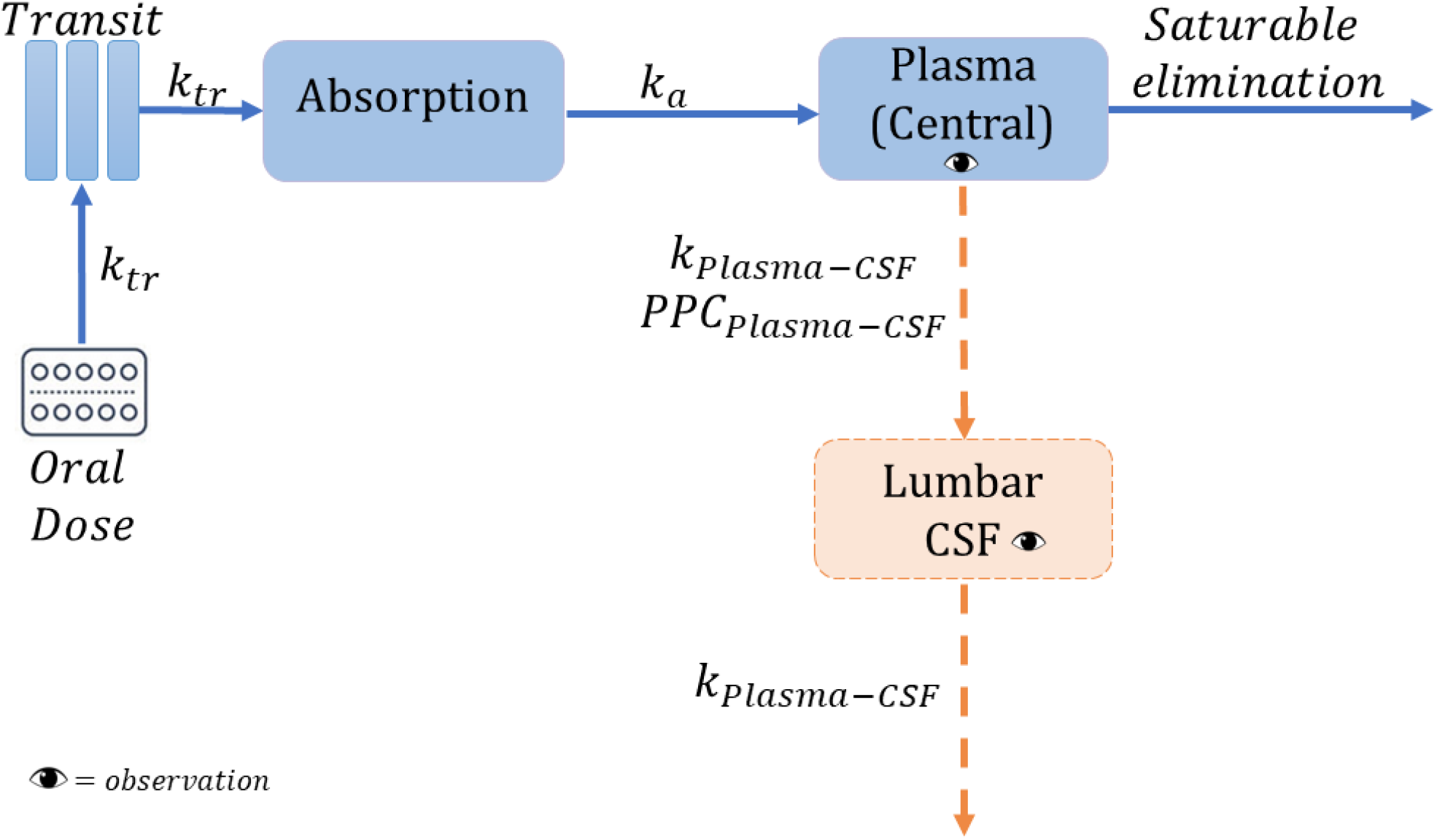
Schematic representation of the final model. *k*_*tr*_ is the rate constant for the drug passage through the transit compartments, *k*_*Plasma*−*CSF*_, equilibration rate constant plasma-cerebrospinal fluid CSF) which describes how soon the change in plasma is reflected in the CSF; *PPC*_*Plasma*−*CSF*_, the pseudo-partition coefficient which represents the ratio of drug in CSF to the plasma.

The CSF concentrations were linked to the plasma concentrations with an equilibration half-life of 3.5 hours (95% confidence interval, 2.13 - 7.33) and the steady-state equilibrium ratio (*PPC*), indicating the relative amount of linezolid exposure in CSF, which was dependent on CSF protein levels. The *PPC*-CSF protein relationship was described using a piece-wise linear (broken-stick) function, where the *PPC* increased with higher CSF protein levels until a maximal CSF protein value where the *PPC* plateaued (i.e., a maximal *PPC* value). The breakpoint was estimated, while the slope (i.e., the change in *PPC* per change in CSF protein) was calculated from the breakpoint and the intercept (minimum *PPC*) which was fixed to be 0 to prevent the estimation of negative values of *PPC* which are physiologically unplausible. For each 0.1 mg/mL increase in CSF protein, we found an increase of 3% in *PPC* up to 1.18 mg/mL of CSF protein, after which the *PPC* reached a maximal value of 36.5% (95% CI, 23.8% – 56.6%) (**Figure 2)**. Both CSF protein and CSF albumin were found to correlate significantly with *PPC*; however, the two are highly positively correlated. Only CSF protein was included in the final model because it resulted in a more significant drop in OFV and because albumin is a component of the proteins measured.

**Figure 2:**
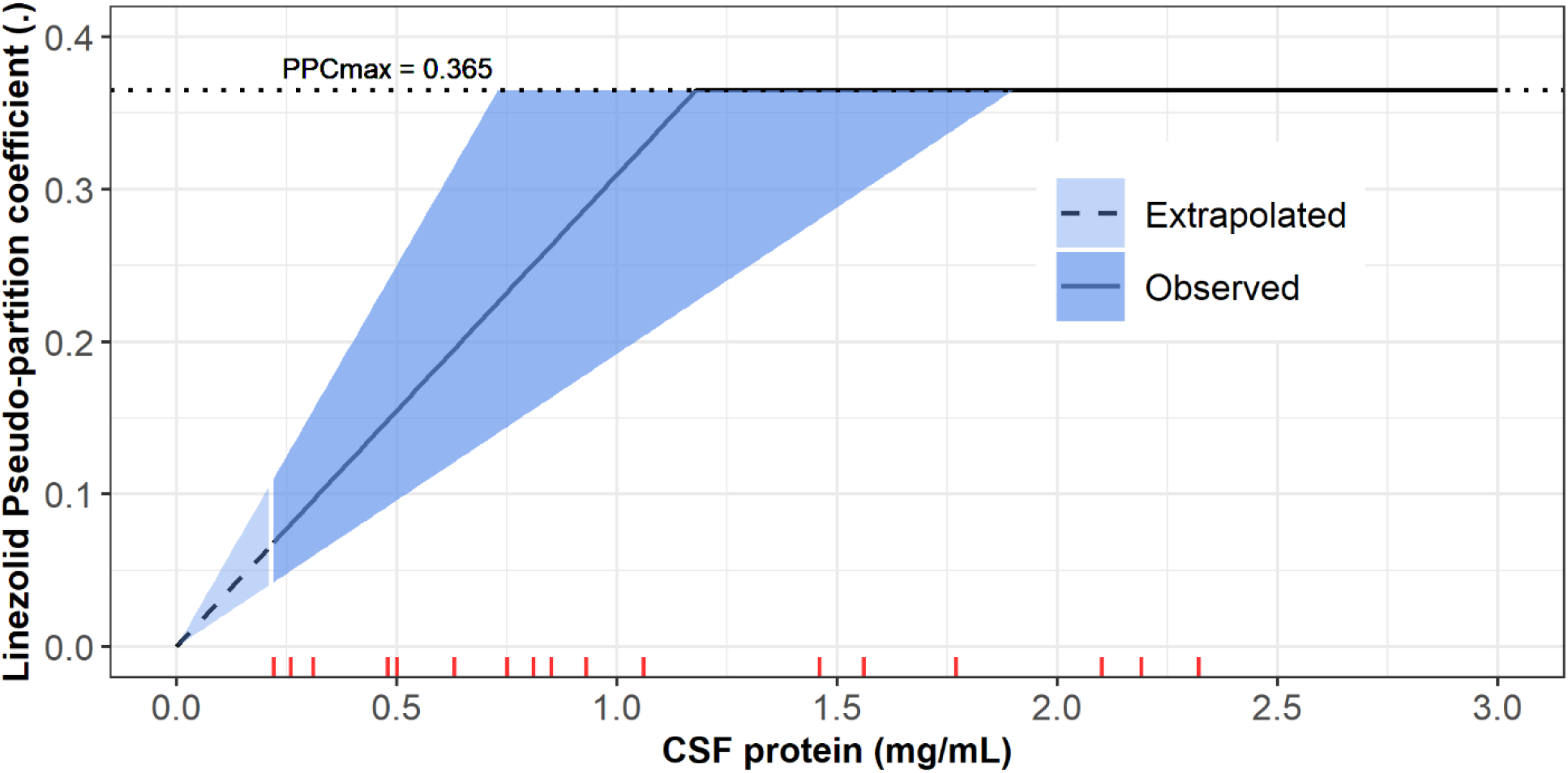
The relationship of *PPC* vs the CSF protein level using the piece-wise (broken-stick) function. The solid line represents the median and the shaded areas represent the uncertainty around the estimates of the breakpoint (the maximal CSF protein value at which *PPCmax* is reached) and the calculated slope. The dashed line depicts the extrapolated part of the *PPC*-CSF protein relationship for CSF protein values outside the range observed in the study cohort (The lowest observed value was 0.22 mg/mL). The red ticks represent the values of CSF protein observed in our cohort (values above 3 were truncated for better figure visibility).

**Figure 3:**
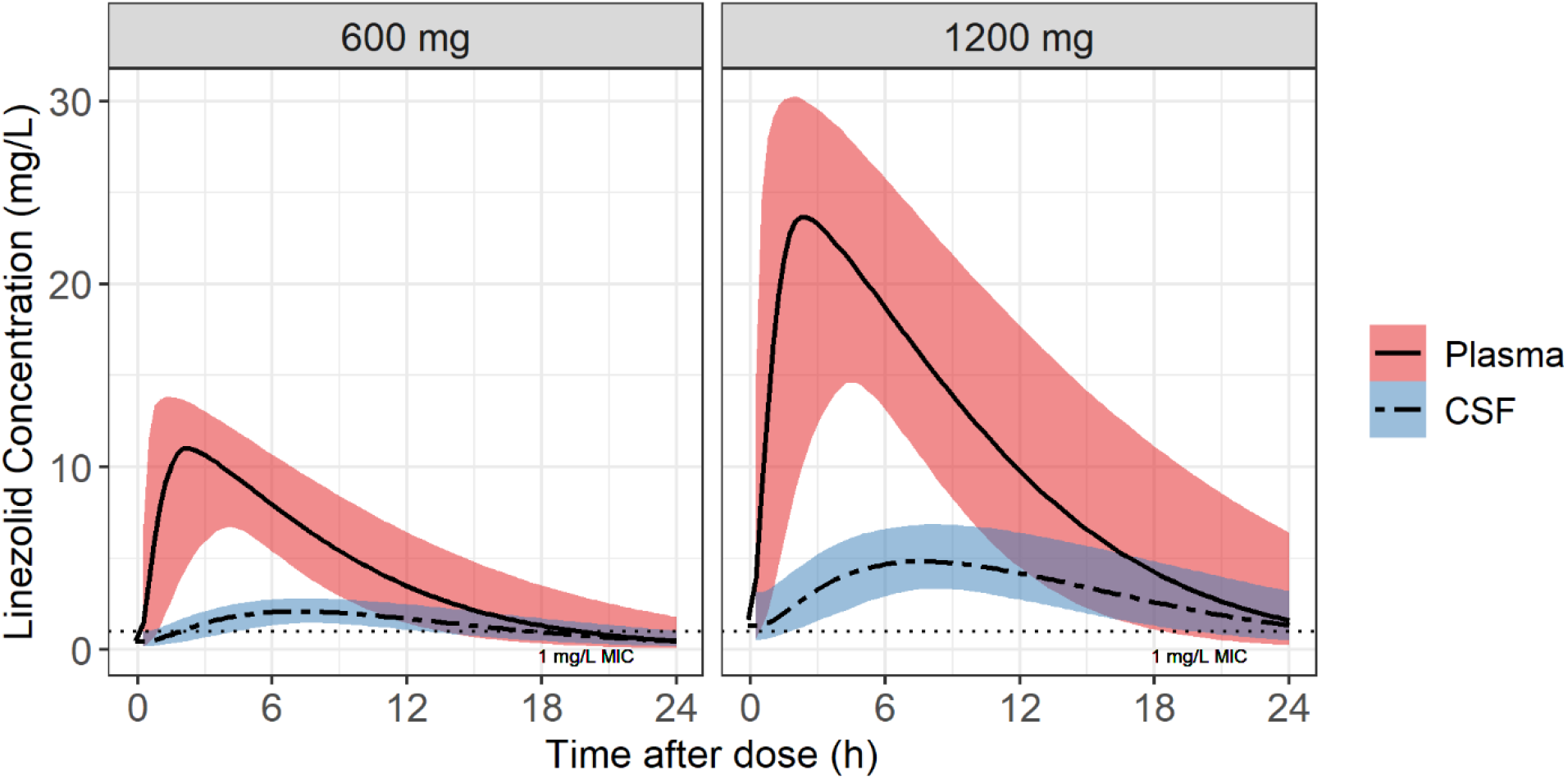
Simulated typical concentration-time profiles for plasma and cerebrospinal fluid (CSF) for the 1200 mg and 600 mg oral daily dose of linezolid. The solid and dashed lines represent the median for the plasma and CSF, respectively and the shaded areas represent the 90% confidence intervals. The horizontal dotted line indicates the wild-type minimum inhibitory concentration (MIC) value of linezolid for *M. tuberculosis*.

**Figure 4:**
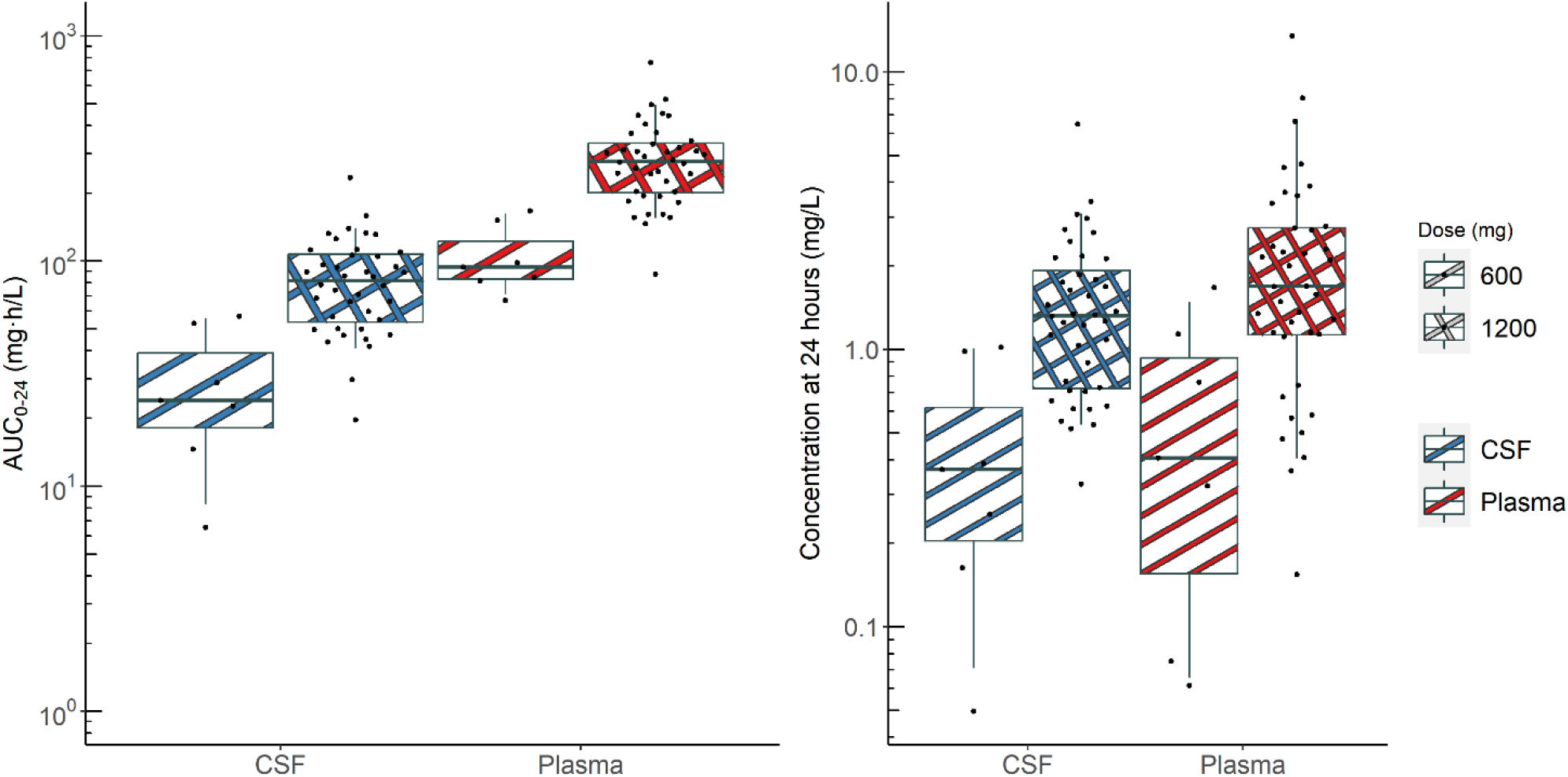
Box and whisker plots showing the secondary model-derived exposure parameters, *AUC*_0−24*h*_ and concentration at 24 hours post-dose (*C*_24*h*_) stratified by dose. The dots represent individual values; whiskers are the 2.5^th^ and 97.5^th^ percentiles (n = 7 for 600 mg and 40 (30 on day 3 plus 10 on day 28) for 1200 mg).

### Simulations

The simulated plasma and CSF concentration time profiles for the typical participant in our cohort following a once daily dose of either 600 mg or 1200 mg linezolid are depicted in **Figure 2** and model-derived individual values for the steady-state AUC_0-24h_ and trough concentrations are summarized in **Table 3**.

**Table 3.**
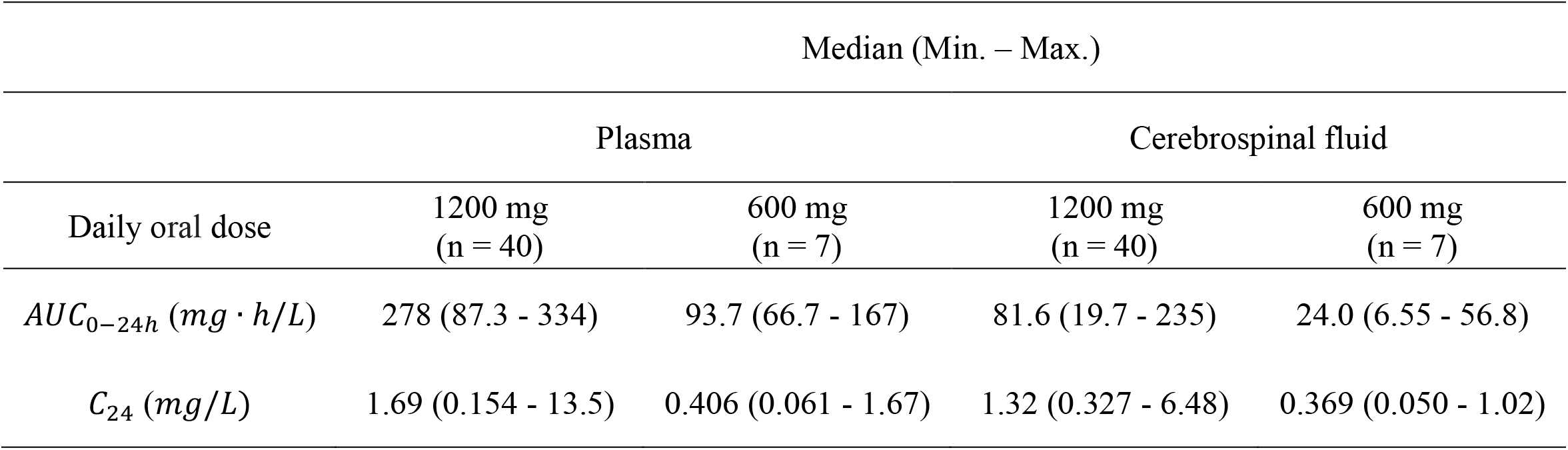
Linezolid model-derived area under the curve for 24 hours and concentrations at 24 hours post-dose

## DISCUSSION

Linezolid is being evaluated in several clinical trials as part of enhanced antimicrobial therapy for TBM. This is based on limited clinical evidence from small observational studies in TBM [10,11] and reports of successful use in gram-positive CNS infection. However, there is scarce information on linezolid exposure in the CSF, especially among patients with TBM, a presumed requirement for clinical efficacy in this condition. We characterized the PK of linezolid in plasma and CSF from a cohort of South African patients with HIV-associated TBM. The extent of linezolid penetration into the CSF was ∼30% on average of plasma exposure and correlated with CSF protein concentrations – CSF penetration was higher in participants with higher CSF protein, reaching a maximal value of ∼37%. Co-administration with high-dose rifampicin (35 mg/kg/day), when comparing the duration on rifampicin treatment on day 3 versus day 28 did not have a significant effect on the PK of linezolid.

Several prior studies may help to contextualize our findings. A recent observational study reported CSF linezolid concentrations from 17 TBM patients (only one with HIV) who received linezolid 600 mg daily [25]. The median CSF concentrations were 0.90 mg/L and 3.14 mg/L and the CSF/serum ratios were 0.25 and 0.59 at 2- and 6-hours post-dose, respectively. CSF linezolid concentrations have also been reported from two small cohorts of neurosurgical patients receiving 600 mg linezolid intravenously every 12 hours. In the smaller study (n = 7) the mean observed CSF-to-plasma AUC ratio was 0.565 (n = 7), the mean (±standard deviation) *AUC*_0−∞_ after the first dose was 37.7 (±23.9) mg·h/L and *AUC*_0−12*h*_ after the fifth dose was 53.7 (±50.3) mg·h/L. In the slightly larger study (n = 14) mean observed CSF-to-plasma AUC ratio was 0.66 and mean (±standard deviation) AUC in CSF was 101 ±59.6 mg.h/L [26,27]. Direct comparison is limited because of differences in population (HIV status, disease type and severity), dosing and administration, and drug assays. CSF/plasma concentration and AUC ratios should be cautiously interpreted in these prior studies [25–27] since observed CSF and plasma concentrations were compared at the same timepoints, not accounting for delay in distribution between the plasma and CSF. Despite having access to only a single CSF sample per visit (due to the invasive nature of lumbar puncture), using a model-based approach allowed us to describe the time course for linezolid entry into CSF. The limitation of sparse CSF sampling in our study was further mitigated by randomizing participants to different sampling times so that CSF samples could be obtained over the full dosing interval.

Other studies have also reported a relationship between the levels of CSF total protein (or albumin) and antituberculosis drugs in TBM [28,29]. In a pediatric population there was a linear relationship between CSF protein concentration and the CSF penetration of rifampicin, with a 63% increase in the penetration coefficient for every 10-fold change in protein levels [28]. In another pediatric TBM cohort, an exponential function was used to describe the relationship between CSF protein concentrations and the partition coefficient of rifampicin where an increase of 1 g/L in CSF protein concentration resulted in a 1.28-fold increase in the partition coefficient [29].

There are two plausible, potentially overlapping, explanations for our finding of a correlation between CSF protein levels and extent of CSF linezolid partitioning. In a healthy state, the BCSFB is intact and only a small fraction of plasma proteins can enter into the CNS, leaving only unbound drug fraction available for penetration into this compartment [7]. Inflammation associated with TB meningitis may increase BCSFB permeability causing both plasma protein and total drug concentrations to be higher in the CSF. Another possible explanation for this relationship is higher endogenous CSF protein production from local inflammation leading to alterations in CSF drug binding kinetics and higher concentrations of total drug in TBM. Quantification of free drug CSF concentrations may help to further delineate CSF protein-drug relationships.

Linezolid is provided with high dose rifampicin (35 mg/kg/day) in ongoing efficacy trials for TBM. Because of prior reports of a drug-drug interaction between rifampicin and linezolid, plus the likelihood of a rifampicin dose effect on metabolizing enzyme activity [30] which could affect the linezolid plasma exposure and hence the CSF exposure, we investigated a potential effect of rifampicin on linezolid PK. In our study, there was no control group of participants who received only linezolid without rifampicin to clearly identify a drug-drug interaction. However, estimated linezolid clearance in our cohort was comparable to that reported from patients receiving linezolid for drug-resistant pulmonary TB without concomitant rifampicin. In addition, since the maximal cytochrome (CYP) P450 induction effect of rifampicin occurs after at least a week [31], we investigated the effect of the duration of rifampicin therapy (rather than rifampicin co-administration as categorical covariate) on linezolid PK, and could not detect any significant trends. Furthermore, we found no relationship between 4β-OHC:cholesterol or 4β-OHC alone (as predictive biomarker of enzyme induction by rifampicin) and linezolid clearance or bioavailability. Our data indicate that even if rifampicin had an effect on linezolid exposures, it is unlikely to be clinically relevant.

In contrast to our findings, other smaller studies among healthy volunteers and non-TB patients have demonstrated a reduction in linezolid exposure when co-administered with rifampicin [14,32–34]. This interaction has been variously attributed to either a large increase in the expression of the CYP3A4 isoenzyme that typically has a small contribution to linezolid clearance [14] or to increased upregulation of linezolid intestinal secretion by rifampicin induction of P-glycoprotein (P-gp) [34]. There is no definitive evidence that linezolid is a substrate of P-gp, plus it is mainly (∼68%) metabolized in the liver via morpholine ring oxidation, which is independent of the CYP450 system, with the remainder excreted unchanged via the kidneys [14].

As reported for pulmonary TB patients, saturable elimination was observed at higher linezolid plasma concentrations, resulting in non-linear PK [35]. Despite subtle differences in Michaelis-Menten elimination kinetics (*km*) our estimates for *CLmax* and *V* are in line with previously published linezolid models [35–40]. Prior models based on data from non-TB [41] and pulmonary TB patients [42], included an empirical inhibition compartment to describe concentration- and time-dependent autoinhibition of elimination. We also tested this approach, but it did not result in a better model fit for our data, and clearance values estimated by these models are similar to ours.

Our analysis had a few limitations. First, the sparse plasma sampling (3 samples) performed during the 2^nd^ PK visit does not allow for robust estimation of the non-linearity in clearance, especially since only 7 participants were on the reduced dose (600 mg). However, the model fit improved significantly (*p*-value < 0.001) when including saturation of clearance with higher concentrations, supporting this conclusion. Secondly, A limitation of the *PPC*-CSF protein relationship in our model is that the minimum *PPC* was fixed to 0, i.e., no CSF protein means no linezolid gets into the CSF in order to prevent the estimation of negative values of *PPC* which are physiologically implausible. However, a CSF protein value of 0 is not plausible in clinical practice. The total protein concentration of CSF varies between 0.2% and 0.5% of the total protein concentration of blood [43]. It is considered that 80% of CSF proteins originate in blood and that CSF proteins are diluted in a molecule-size-dependent concentration gradient [44]. Thirdly, there was high variability in the observed CSF concentrations (driven by the large proportion of undetected concentrations 25%) which was reflected in estimation of the proportional error for the CSF observations (91.5%). Finally, we did not undertake simulations to estimate probability of target attainment. While our simulations do suggest that 1200 mg daily dosing will achieve linezolid concentrations above the critical concentration MIC of 1 mg/L for *M. tuberculosis*, it is important to note that a PK efficacy target is not established for TBM and that drug protein binding (and relative free fraction of active drug) in the CSF is unknown.

In conclusion, we successfully developed a population PK model for linezolid among adults with HIV-associated TBM, demonstrating that CSF concentrations of linezolid are around 30% of those in plasma, even with concomitant use of high-dose rifampicin. These findings support continued clinical evaluation of linezolid together with rifamycins for the treatment of TBM in adults. Our model provides a platform that can be used for exploring alternative linezolid dosing strategies in TBM once treatment targets are established.

## Data Availability

The data that support this research can be made available upon reasonable request to bona fide researchers by contacting Paolo Denti.

## ACKNOWLEDGMENTS

Computations were performed using facilities provided by the University of Cape Town’s ICTS High Performance Computing team: https://ucthpc.uct.ac.za/. Study data were collected and managed using REDCap (Research Electronic Data Capture) tools hosted at the University of Cape Town. REDCap is a secure, web-based software platform designed to support data capture for research studies [45,46].

