## Supplementary Material for "Linezolid population pharmacokinetic model in plasma and cerebrospinal fluid among patients with tuberculosis meningitis"

### Supplementary File

### Laboratory assays

#### Linezolid

The CSF samples were processed with a protein precipitation extraction method using linezolid-d3 as the internal standard, followed by high-performance liquid chromatography with tandem mass spectrometry detection on a SCIEX API 3200 instrument. The analyte and internal standard were monitored at mass transitions of the protonated precursor ions 338.2 and 341.1 to the product ions 296.3 and 297.3 for linezolid and linezolid-d3, respectively. The calibration curve fitted a quadratic regression (weighted by 1/x) over the range of 0.1 to 20 mg/L. The accuracy of the quality control samples during sample analysis was between 103.5 and 105.5%, with precision of less than 2.1%.

#### 4-beta hydroxy cholesterol (4β-OHC)

4β-OHC was measured with a high-performance liquid chromatography with tandem mass spectrometry assay in the Division of Clinical Pharmacology at the University of Cape Town. The extraction process involved a liquid-liquid extraction, which uses alkaline hydrolysis using potassium hydroxide and chemical derivatization using picolinic acid. Stable isotope labelled 4β-hydroxy cholesterol-d7 (4β-OHC-d7) was used to prepare calibration standards and quality control samples in human plasma. Endogenous 4β-OHC was measured using the surrogate analyte, 4β-OHC-d7. 4β-OHC -d4 was used as the internal standard. Chromatographic separation was done with gradient elution on a Gemini C6 Phenyl analytical column. A Sciex 5500 mass spectrometer at unit resolution in the multiple reaction monitoring acquisition mode was used to monitor the transition of protonated ions to their respective product ions. Electrospray ionization in the positive mode was used for ion production. The calibration curve fitted a quadratic regression (weighted by $1/x^2$) over the range of 2.00 to 500 ng/mL.

### Pharmacokinetic modelling

Nonlinear mixed-effects modelling in NONMEM® 7.5 with first-order conditional estimation with eta-epsilon interaction (FOCE-I) was used to develop a population pharmacokinetic model that describes linezolid pharmacokinetics (PK) in both plasma and lumbar cerebrospinal fluid (CSF). Pirana 3.0.0 software was used for model management; Perl-speaks-NONMEM® (PsN) 4.9.0 and R 4.0.4 via RStudio were used for post-processing NONMEM® results and generating figures[1]. For the plasma model, the nonlinearity in clearance observed at higher doses was accounted for by a concentration-dependent $CL$ described by the following equation:

$$Vmax = CLmax.km$$

where $Vmax$ is the maximal elimination rate in mg/h, $km$ is the linezolid plasma concentration ($Cp$) at which the elimination is half-maximal in mg/L, and $CLmax$ is the maximal clearance reached with increasing $Cp$ in L/h.

Between-subject, between-visit, and between-occasion variabilities were tested for the different plasma and CSF parameters. Each PK sampling day (day 3 and day 28) was considered as a separate visit. Each dose and its following samples were considered a separate occasion, therefore, the dose before the sampling visit along with the predose concentration were treated as a separate occasion from the dose administered during the PK visit and the following concentrations. Residual unexplained variability was described using a combined proportional and additive error model, with the additive error for all samples set to be at least 20% of the LLOQ. Concentrations below the lower limit of quantification (BLQ) were censored according to Beal's M6 method, in which the last censored value in a series during the absorption phase and the first censored value in a series in the terminal phase were replaced with LLOQ/2 and the other censored values in a series were discarded [2]. To account for the larger level of uncertainty in the imputed censored values, their additive error was inflated by LLOQ/2.

The process of model development and covariate inclusion was guided by physiological plausibility, model fit diagnostics, and the drop in the objective function value (OFV). The likelihood ratio test for the drop in OFV was used to compare between nested models, assumed to be approximately $\chi^2$ distributed with n degrees of freedom, where n is the number of additional estimated parameters. A *p*-value of 0.05 was generally used for inclusion and 0.01 for retention. Model performance was evaluated by means of visual predictive checks (VPC). The VPC for the final model stratified into plasma and CSF concentrations is shown in **Figure S1**. Final parameters precision (95% confidence intervals) was obtained by sampling importance resampling (SIR)[3].

### Imputation of missing covariates

Missing covariates such as CSF protein, CSF albumin, and CSF glucose levels were imputed by the median. A different approach was used for the missing heights (necessary for fat-free mass calculation) since it was missing in 60% of the participants. Missing heights were imputed using multiple linear regression as suggested by Johansson and Karlsson[4]. In the first step, participant characteristics, namely sex, weight, and height from a study in a similar population[5] were used to develop a multiple linear regression model for height versus weight by sex and accounting for residual variability in heights. Secondly, this multiple linear regression model was used to estimate the missing heights in NONMEM using a random effect as shown in the equations below:

$$Height_{male,\,i} = (1.53 + \ 0.00281.Weight).e^{0.00133}$$

$$Height_{female,j} = (1.51 + \ 0.00133.Weight).e^{0.00215}$$

where height and weight are in m and kg, respectively.

### Effect compartment modelling for CSF concentrations

The CSF concentrations were modelled as dependent on plasma concentrations using an effect compartment, as previously proposed and implemented by Sheiner et al. and Savic et al.[6,7]. Effect compartments are assumed to have a negligible volume compared to the central compartment, with negligible drug transfer between the two compartments. The following differential equation summarizes the kinetics of the effect compartment:

$$\frac{dC_{CSF}}{dt} = k_{plasma-CSF} \cdot \left(PPC \cdot C_{plasma} - C_{CSF}\right),$$

where $k_{plasma-Eff}$ is the first-order equilibration rate constant of the drug between the central compartment (i.e., plasma) and the effect compartment (i.e., CSF), $PPC$ is the pseudo-partition coefficient, $C_{plasma}$ and $C_{CSF}$ are the drug concentration at time $t$ in plasma or CSF, respectively. Figure S2 shows the interpretability of the equilibration rate constant and the PPC in the context of effect compartment modelling approach.

### Figures

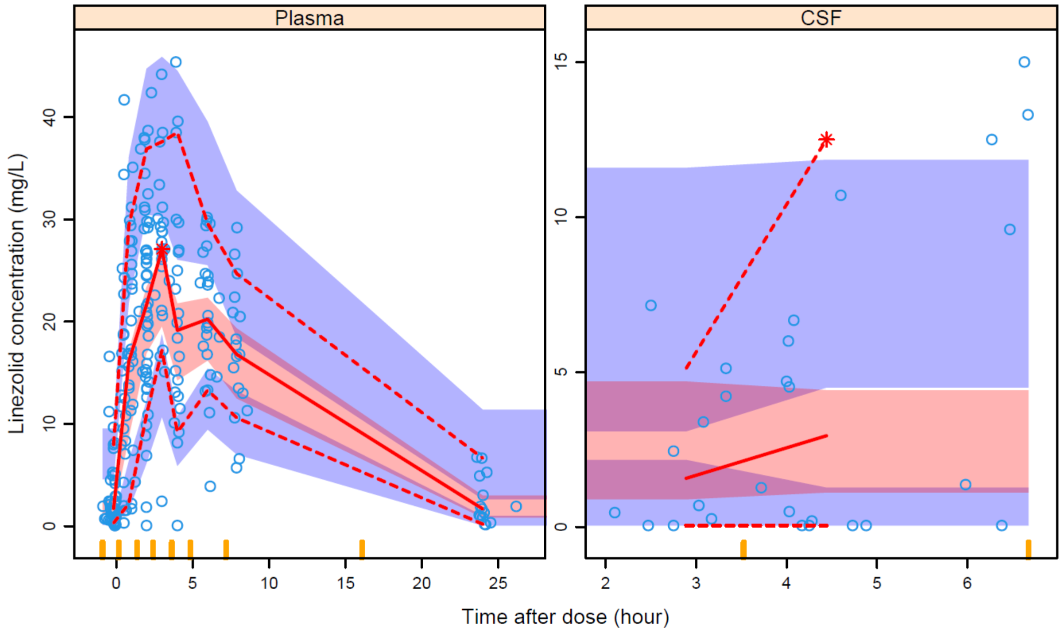


**Figure S1:** Visual predictive check (VPC) (n=1000) showing plasma drug concentration versus time after dose for the final models stratified into plasma and cerebrospinal fluid. The dots are the original observations; the solid line is the median and the dashed lines are the 10th and 90th percentiles of the observed data; the shaded areas are the 95% confidence intervals of the same percentiles as simulated by the model. A suitably fitting model will have most of the observed percentiles within the simulated confidence intervals.

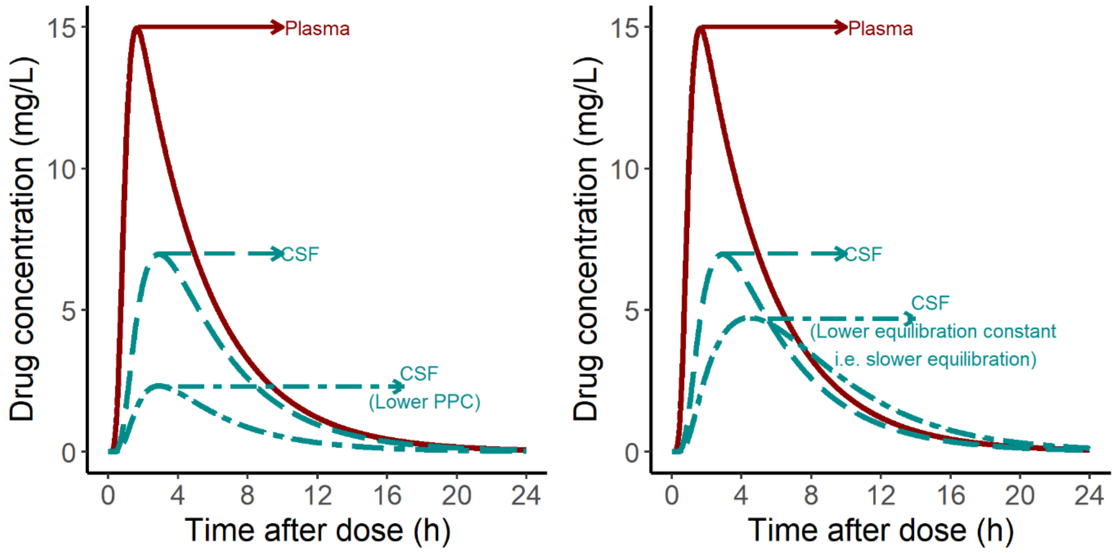


**Figure S2:** Demonstration on the interpretability of the equilibration rate constant and the PPC in the context of effect compartment modeling approach.
